# Assessing the risk of endogeneity bias in health and mortality inequalities research using composite measures of multiple deprivation which include health-related indicators: A case study using the Scottish Index of Multiple Deprivation and population health and mortality data

**DOI:** 10.1101/2022.12.01.22282961

**Authors:** D R R Bradford, M Allik, A D McMahon, D Brown

## Abstract

The inclusion of health-related indicators in composite measures of multiple deprivation introduces a risk of endogeneity bias when using the latter in health inequalities research. This bias may ultimately result in the inappropriate allocation of healthcare resources and maintenance of preventable health inequalities. Mitigation strategies to avoid this bias include removing the health-related indicators or using single constituent domains (such as income or employment class) in isolation. These strategies have not been widely validated. This study used population-level health and mortality data with a contemporary composite measure of multiple deprivation (Scottish Index of Multiple Deprivation; SIMD) to assess these mitigation strategies. The differences between deprivation methods (original, health excluded, and income domain) were negligible. The results of quantitative research on health inequalities are unlikely to be affected by endogeneity bias.

## 1. Introduction

### 1.1. Background

Deprivation is a multi-faceted state which manifests in many aspects of people’s lives and is affected by more than just monetary and material factors (Braveman et al., 2005; Liberatos et al., 1988; Townsend, 1987; Watson et al., 2019). People living in poor socioeconomic circumstances face significantly worse outcomes for morbidity and mortality than those living in areas of lesser deprivation (Agardh et al., 2011; Authors, 2019a, 2020; Frølich et al., 2019; Knies and Kumari, 2022; Mackenbach et al., 2008; Schiøtz et al., 2017). Developing policy which addresses inequalities in morbidity and mortality requires accurate measures of deprivation, which are commonly implemented at the area level. Measures of area-level deprivation must, therefore, be implemented both methodologically and theoretically appropriately to avoid the maintenance or worsening of preventable inequalities in health.

Contemporary area-level composite measures of multiple deprivation (CMMDs) attempt to account for the multidimensional nature of deprivation. They go beyond typical single-item measures such as household income or socioeconomic employment categories by including indicators of different forms of deprivation. This increases the accuracy and robustness with which the latent construct of deprivation is measured (Bryere et al., 2017; Lian et al., 2016; Strömberg et al., 2021). Some CMMDs incorporate indicators based on health-related manifestations of deprivation, including CMMDs used in the UK (McLennan et al., 2019; Northern Ireland Statistics and Research Agency, 2017; Scottish Government, 2020; Welsh Government, 2019), New Zealand (Exeter et al., 2017), Australia (Australian Bureau of Statistics, 2018), and Germany (Maier et al., 2013). However, it has been suggested that the inclusion of health-related indicators in the construction of CMMDs could, theoretically, introduce endogeneity bias when used alongside socioeconomically-graded health outcome variables. This study explores the potential effects of this bias on the results of quantitative health inequalities analysis.

### 1.2. Source, consequences, and mitigation of endogeneity bias in composite measures of multiple deprivation

Endogeneity bias can occur when outcome and predictor variables have a direct relationship (Archie, 1981; Hill et al., 2021). For example, analyses that examine differences in mortality rates by area-level deprivation can be biased if the chosen CMMD incorporates mortality rates as an indicator (i.e., simultaneity bias; Hill et al., 2021). Alternatively, the association between predictor and outcome variables may not be explicit. But if both are strongly influenced by a latent variable, such as health-related deprivation, then bias can still occur. Conceptually, the circular reasoning being expressed in such cases is that a deprivation-dependent health outcome is partially specified to be a function of deprivation-dependent health (through the inclusion of health-related indicators in the CMMD). The potential consequences of this endogenous link between health-related outcome and predictor variables have not been given sufficient attention in the research literature.

The potential for results in socioeconomic inequalities health research to be biased should be a substantial concern for researchers, policy makers, and the public. With the move toward evidence-based public health decision making, the influence of health inequality research on policy making is growing. These decisions dictate where health and healthcare resources are directed (Baczyk et al., 2016; NHS England and Improvement, 2021; Salmond and Crampton, 2012). Erroneous results caused by endogeneity bias can, therefore, potentially lead to a failure to address preventable socioeconomic inequalities in morbidity and mortality.

The risk posed by the consequences of endogeneity bias is recognised by researchers in both academia and government. Official guidance from the Scottish and British governments recommends that analyses featuring health or mortality outcomes should either 1) exclude the health domain from their chosen CMMD and recalculate deprivation scores, or 2) use the income domain only (known to correlate strongly with overall deprivation; McLennan et al., 2019; Ralston et al., 2014; Scottish Government, 2022). Health inequality researchers looking to mitigate this risk may choose one of these two methods, or explicitly note the potential for endogeneity bias as a limitation of their work (e.g., Authors, 2009, 2016, 2022; Fenton et al., 2019; Jordan, 2004; Ralston et al., 2014). Although using these mitigation strategies to assign deprivation scores can be appropriate — and researchers should certainly make informed judgements when selecting a measure of deprivation — formal validation or justification of these alternative methods of assigning deprivation scores is rare. The purpose of the present study is to provide formalised evidence of the effects of such strategies and explore how it may affect the results of health inequality research.

To the best of our knowledge, the only published examination of the effect of endogeneity bias between health outcomes and the health domain of a CMMD was reported by Adams and White (2006). The authors calculated deprivation scores for the 2004 Index of Multiple Deprivation (IMD) for England after removing the health domain. Their results showed no significant differences between analyses which used either the original all-domain or the health-excluded version of the IMD. The lack of evidence around this topic is further highlighted by the absence of studies which consider the effects of endogeneity when mortality is the outcome variable. This is of particular importance since CMMDs can include mortality-related indicators in the health domain and, again, this has previously been recognised as an issue by researchers (e.g., Fenton et al., 2019).

### 1.3. Scotland as a case study

Scotland was selected as a case study to investigate concerns about endogeneity bias in the context of CMMD and health inequalities analysis. It was chosen as 1) it experiences significant health inequalities which are worse than comparably developed countries (McCartney et al., 2012), 2) there are high-quality data available about population-level health outcomes and mortality, and 3) it has a widely-used area-level CMMD which includes health-related indicators of deprivation: the Scottish Index of Multiple Deprivation (Scottish Government, 2020).

### 1.4. Study aim and objectives

The aim of this study was to assess whether the endogeneity bias described above can have a substantial effect on the results of exemplary health inequalities analyses. This work was intended to expand on the results of Adams and White (2006) by adding a new context (Scotland), an additional outcome (mortality), and testing another common bias mitigation strategy (using the income domain in isolation). Moreover, by testing the effect of common bias-mitigation strategies used in health inequalities research, this work aimed to provided evidence to help researchers justify or reject these strategies.

The first objective of this study was to assess concordance between overall SIMD deprivation scores and the deprivation scores arising from the two alternative methods of calculating deprivation scores described above: 1) excluding the health domain indicators from the CMMD, and 2) using the income domain in isolation. The second objective was to report how the results of analyses which use population-level health and mortality data in Scotland were affected by using these two alternative deprivation calculation methods.

## 2. Data and methods

### 2.1. Composite measure of multiple deprivation

The Scottish Index of Multiple Deprivation (SIMD) provides an estimate of the combined extent of deprivation in multiple domains, homogenised over a small geographic area known as a data zone (described in more detail below; Scottish Government, 2020). An overall SIMD score is calculated for each data zone using area-level indicators of deprivation across the following domains, with the relative weight of each domain shown in parentheses: income (0.28); employment (0.28); education, skills, and training (0.14); geographic access to services (0.09); crime (0.09); housing (0.02); and health (0.14). The health domain of the SIMD is constructed from seven weighted indicators detailed in Supplementary Table S1.

Individual data zones can be ranked by their composite deprivation score or their score in each constituent domain. Ranks are used rather than raw scores because the latter are not meaningful when comparing across domains. This is due to heterogeneity among indicator scoring methods. SIMD ranks are published for each data zone by the Scottish Government, along with technical guidance about relevant calculation methods (http://simd.scotland.gov.uk/). For this study we compared three methods of calculating data zone deprivation ranks: 1) overall SIMD rank including the health domain, 2) overall SIMD rank excluding the health domain (hereafter referred to as the Scottish Index of Multiple Deprivation Excluding Health; SIMDEH), and 3) the income domain in isolation.

SIMD deprivation ranks are most commonly used to categorise data zones into population-weighted deprivation quantiles (e.g., Authors, 2018, 2021; Covvey et al., 2014; Henery et al., 2021; Thompson et al., 2013). For example, the data zones which, after ranking by deprivation, cumulatively contain 10% of the total population in the most deprived areas are assigned to deprivation decile 1 in the SIMD ordering convention. The least deprived data zones containing 10% of the total population are assigned to decile 10. In this study we examined quintiles, deciles, and vigintiles. For brevity, only the results of analyses which used deciles are reported here. It should be noted that no substantial differences with the results presented were identified using different numbers of quantiles.

The 2012 and 2020 iterations of the SIMD were used in the present study. *SIMD2012* divides Scotland into 6505 data zones with a mean population of 803 (standard deviation = 248). This iteration of the SIMD was used when analysing health data from the 2011 Scottish Census and 2010–2012 mortality data (detailed below). *SIMD2020v2* comprises 6796 data zones with a mean population of 778 (standard deviation = 219). This iteration of the SIMD was used during the analysis of 2017–2019 mortality data.

### 2.2. Self-rated health

Data on self-rated health were available from the 2011 Scottish Census Commissioned Table CT_0033d_2011 (National Records of Scotland). This provided population-level data on census respondents’ perception of their own health, the health of other members of their household, and the health of their dependants. The census question was phrased “*How is your health in general?*” with possible responses being *very good, good, fair, bad*, and *very bad*. Response counts were available separately for males and females for each data zone in five-year age groups from ages 0–74, with all individuals aged 75 years or over being placed into a single age group. A *bad health* count variable was constructed by combining *bad* and *very bad* responses. The denominator used in self-rated health analyses was the sum of the data zone populations extracted from the 2011 Scottish Census Commissioned Table.

### 2.3. Limiting long-term health conditions

Data on the number of individuals with any self-reported limiting long-term health conditions (LLTHC) were available from the 2011 Scottish Census Commissioned Table CT_0033f_2011 (National Records of Scotland). The census question was phrased “*Do you have any of the following conditions which have lasted, or are expected to last, at least 12 months?*” Respondents could select from a predefined list of common LLTHC such as diabetes or severe visual impairment, or they could provide a free text response. Response counts were available separately for males and females for each data zone in five-year age groups from ages 0–74, with all individuals aged 75 years or over being placed into a single age group. The responses to this question were used to construct a count of how many individuals had a LLTHC. The denominator used in LLTHC analyses was the sum of the data zone populations extracted from the 2011 Scottish Census Commissioned Table.

### 2.4. Mortality

Mortality counts were available from the National Records of Scotland. Mortality data were available separately for males and females for each data zone in five-year age groups from ages 0–74. All individuals aged 75 years or over were placed into a single age group to align with the structure of the census health outcomes data. Counts were obtained for 2010–2012 (inclusive) and averaged, referred to as 2011 data hereafter. These years were selected to be closest in time to the health data from 2011 Census. Mortality data for 2017–2019 were similarly averaged, referred to as 2018 data hereafter. The 2017–2019 data were selected as the most recent mortality data available which are not affected by the excess deaths associated with the COVID-19 pandemic. Multiple years of mortality counts were averaged to counter the annual variability in death counts. It should be noted that mortality (and all other outcomes) were aggregated across all data zones within a deprivation decile. As such, there is no concern about error due to very low or zero counts which may have occurred in some data zones. The denominator for 2011 mortality analyses was the sum of the data zone populations extracted from the 2011 Scottish Census Commissioned Tables. The denominator for the 2018 mortality analyses was the mean of the 2017–2019 mid-year population estimates (Public Health Scotland).

### 2.5. Data statement

Mortality data are available from National Records of Scotland on request. Census data and population estimates are publicly available from National Records of Scotland (National Records of Scotland) and Public Health Scotland (Public Health Scotland), respectively.

### 2.6. Analysis methods

Health outcome, mortality, and deprivation decile data were linked for each data zone. Data zones were excluded from all analyses if they had zero population. This led to the exclusion of five data zones from the 2011 analyses and three from the 2018 analyses. Confidence intervals were set at 95% throughout. Analyses were carried out in R v.4.1.3 (R Core Team, 2021) and employed functions from the *Hmisc* (Harrell Jr, 2021), *PHEindicatormethods* (Georgina, 2020), *psych* (Revelle, 2022), *SocEpi* (Authors), and *tidyverse* (Wickham et al., 2019) packages.

The strength of concordance was assessed between the overall SIMD and, independently, the two alternative deprivation methods under investigation (SIMDEH and income domain). The correlation of the deprivation ranks between deprivation methods was assessed using Spearman’s rank correlation coefficient (*r*_*s*_) and visualised with Bland–Altman plots. Crude proportion of agreement and Cohen’s kappa (*K*_*W*_ ; Cohen, 1968) were used to assess the concordance of assigned quantiles between deprivation methods. *K*_*W*_ was applied using equal weighting for each level of quantile discordance.

Health and mortality outcomes were analysed separately for males and females throughout. The prevalence of health outcomes and mortality incidence were age standardised at the decile level to the 2013 European Standard Population (Eurostat, 2013). Standardisation was used to adjust for differences in age distributions between deciles (National Records of Scotland, 2022). Standardisation was carried out for both the overall population and select age subgroups (0–19, 20–44, 45–74, 75+ years; subgroup results not shown but were essentially similar to the overall population results). Standardised rates were calculated using the *PHEindicatormethods* package for R (Georgina, 2020).

Absolute inequalities were assessed using the Slope Index of Inequality (SII). In the context of the present work, the SII indicates the absolute difference in the age-standardised rate of people experiencing a health outcome between the notionally most and least deprived data zones (Authors, 2019c; Regidor, 2004; Scottish Public Health Observatory). The SII is therefore a useful measure for informing healthcare resource allocation to address health inequities. Positive values indicate worse outcomes in more deprived areas. SII values were calculated using ordinary least squares regression fitted to age-standardised prevalence rates or mortality incidence rates at the midpoints of deciles using the SocEpi package for R (Authors).

Relative inequalities were assessed using the linear Relative Index of Inequality (RII). The RII is defined here as the SII divided by the relevant age-standardised prevalence or incidence rate for the overall population (Authors, 2019c; Pamuk, 1985; Regidor, 2004; Scottish Public Health Observatory). The RII complements the SII by indicating the degree of association between deprivation and a health outcome. RII values typically lie in the range -2 to 2. However, the magnitude of RII can be greater than 2 if non-linearity is present in the relationship between the health outcome and deprivation data used in the regression model which generates SII values (Authors, 2019c). An RII value of zero indicates no inequality. Positive values indicate worse outcomes in more deprived areas. RII values were calculated using the SocEpi package for R (Authors).

## 3. Results

### 3.1. Deprivation rank and quantile concordance

Pairwise agreement statistics between overall SIMD and both the SIMDEH and income-only deprivation methods are reported in Table 1. The alternative methods led to some data zones being assigned to higher or lower quantiles. Increasing the number of quantiles naturally led to greater numbers of data zones being reassigned to those higher or lower quantile levels. This explains the decreasing value of crude agreement percentage with increasing number of quantiles (see Supplementary Table S4 for a more detailed breakdown of these shifts). However, the more nuanced concordance statistics (*r*_*s*_ and *K*_*W*_) suggested very strong or near-perfect pairwise agreement between SIMD and the two alternative deprivation methods. Bland–Altman plots showed that SIMDEH deprivation ranks for data zones were generally closer to original SIMD ranks than income domain ranks (see Figures S1–S4).

**Table 1.** Concordance statistics between the overall Scottish Index of Multiple Deprivation (SIMD) and both the SIMD Excluding Health (SIMDEH) and income domain of the SIMD (with 95% CI’s in parentheses where available). Presented for the 2012 and 202v2 iterations of the SIMD. *r*_*s*_ = Spearman’s rank correlation coefficient, *%* = crude agreement percentage, *K*_*W*_ = Cohen’s weighted Kappa.

|  | SIMD2012 |  | SIMD2020v2 |  |
| --- | --- | --- | --- | --- |
|  | SIMDEH | Income | SIMDEH | Income |
| *Ranks* |  |  |  |  |
| $r_s$ | 1.00 | .97 | 1.00 | .97 |
| *Quintiles* |  |  |  |  |
| % | 92.0 (91.3, 92.6) | 75.7 (74.6, 76.7) | 93.3 (92.7, 93.9) | 76.6 (75.6, 77.6) |
| $K_W$ | 0.95 (0.91, 0.99) | 0.85 (0.81, 0.88) | 0.96 (0.92, 1.00) | 0.85 (0.82, 0.89) |
| *Deciles* |  |  |  |  |
| % | 82.6 (81.6, 83.5) | 53.1 (51.8, 54.3) | 86.5 (85.7, 87.3) | 56.9 (55.8, 58.1) |
| $K_W$ | 0.95 (0.94, 0.95) | 0.84 (0.83, 0.84) | 0.96 (0.95, 0.97) | 0.85 (0.85, 0.86) |
| *Vigintiles* |  |  |  |  |
| % | 65.8 (64.7, 67.0) | 33.1 (31.9, 34.2) | 72.5 (71.5, 73.6) | 36.4 (35.2, 37.5) |
| $K_W$ | 0.95 (0.94, 0.95) | 0.84 (0.83, 0.84) | 0.96 (0.96, 0.96) | 0.85 (0.85, 0.85) |

### 3.2. Health outcomes

Descriptive statistics for health outcomes are reported in Table 2. There was no meaningful difference between males and females in the proportion of individuals in poor health. This was consistent across age subgroups and SIMD deciles (results not shown). Standardised rates for health outcomes also exhibited no difference between males and females (Table 3).

**Table 2.**
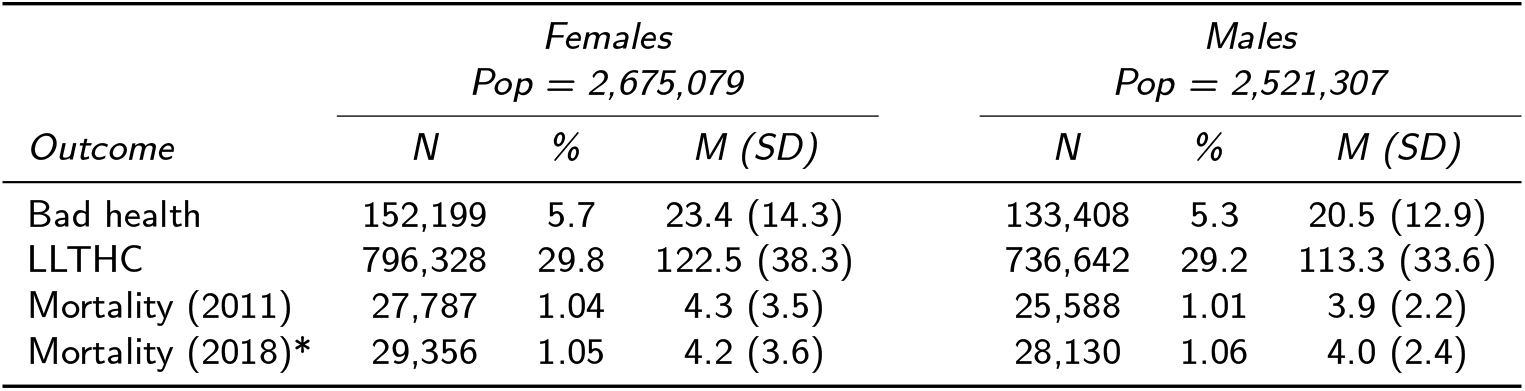
Health and mortality outcome statistics. LLTHC = limiting long-term health condition, *N* = total count of individuals experiencing the health outcome or three-year averaged death count, *%* = percentage of population, *M (SD)* = mean count and standard deviation per data zone. *\** Populations for 2011 shown in header row, populations for 2018: female = 2,791,382, male = 2,650,685.

| Outcome | Females<br>Pop = 2,675,079 |  |  | Males<br>Pop = 2,521,307 |  |  |
| --- | --- | --- | --- | --- | --- | --- |
|  | N | % | M (SD) | N | % | M (SD) |
| Bad health | 152,199 | 5.7 | 23.4 (14.3) | 133,408 | 5.3 | 20.5 (12.9) |
| LLTHC | 796,328 | 29.8 | 122.5 (38.3) | 736,642 | 29.2 | 113.3 (33.6) |
| Mortality (2011) | 27,787 | 1.04 | 4.3 (3.5) | 25,588 | 1.01 | 3.9 (2.2) |
| Mortality (2018)* | 29,356 | 1.05 | 4.2 (3.6) | 28,130 | 1.06 | 4.0 (2.4) |

**Table 3.** Age-standardised health and mortality outcome statistics. ASR = overall age-standardised prevalence rate per 1,000 population for health outcomes and age-standardised incidence rate per 100,000 population for mortality, SII = Slope Index of Inequality, RII = Relative Index of Inequality (95% confidence intervals in parentheses). SII and RII calculated using overall Scottish Index of Multiple Deprivation deciles are presented here but differences in point estimates were negligible when compared with those calculated using Scottish Index of Multiple Deprivation Excluding Health and income-only deprivation methods.

| Outcome | ASR | SII | RII |
| --- | --- | --- | --- |
| Females |  |  |  |
| Bad health | 57.9 (57.6, 58.2) | 97.0 (96.1, 98.0) | 1.67 (1.66, 1.69) |
| LLTHC | 302.5 (301.9, 303.2) | 155.0 (152.8, 157.3) | 0.51 (0.50, 0.52) |
| Mortality (2011) | 1,095 (1,082, 1,108) | 586 (538, 632) | 0.54 (0.49, 0.58) |
| Mortality (2018) | 997 (986, 1,009) | 707 (668, 743) | 0.71 (0.67, 0.75) |
| Males |  |  |  |
| Bad health | 58.1 (57.8, 58.4) | 103.8 (102.8, 104.8) | 1.79 (1.77, 1.81) |
| LLTHC | 315.7 (315.0, 316.5) | 152.2 (149.5, 155.0) | 0.48 (0.47, 0.49) |
| Mortality (2011) | 1,324 (1,307, 1,340) | 1,051 (994, 1,112) | 0.79 (0.75, 0.83) |
| Mortality (2018) | 1,225 (1,211, 1,239) | 1,078 (1,019, 1,127) | 0.88 (0.84, 0.92) |

Increased deprivation was associated with poorer self-reported health outcomes. This was true for both males and females, and across age subgroups (0–19, 20-44, 45-74, 75+ years; results not shown). This trend is exemplified in Figure 1 which presents rates of self-rated bad health in females across deprivation deciles. Results were essentially similar in males. Importantly, this plot demonstrates the overlap in calculated rates between the three deprivation methods. Differences in the age-standardised rates resulting from the overall SIMD and alternative deprivation calculation methods were statistically insignificant in all but two of the 160 pairwise combinations of sex, outcome, and decile. One of these cases was when considering mortality in females in decile 9 where using the original SIMD generated an estimate of 943 per 100,000 population (95% CI: 904, 983) compared with 854 (95% CI: 816, 893) using the income domain. The other case was for bad health in males in decile 6 where using the original SIMD generated an estimate of 45.8 per 1,000 population (95%CI: 45.0, 46.7) compared with 47.6 (95%CI: 46.8, 48.6) using the income domain. However, the magnitude of these within-decile differences were insubstantial compared to across-decile differences.

**Figure 1:**
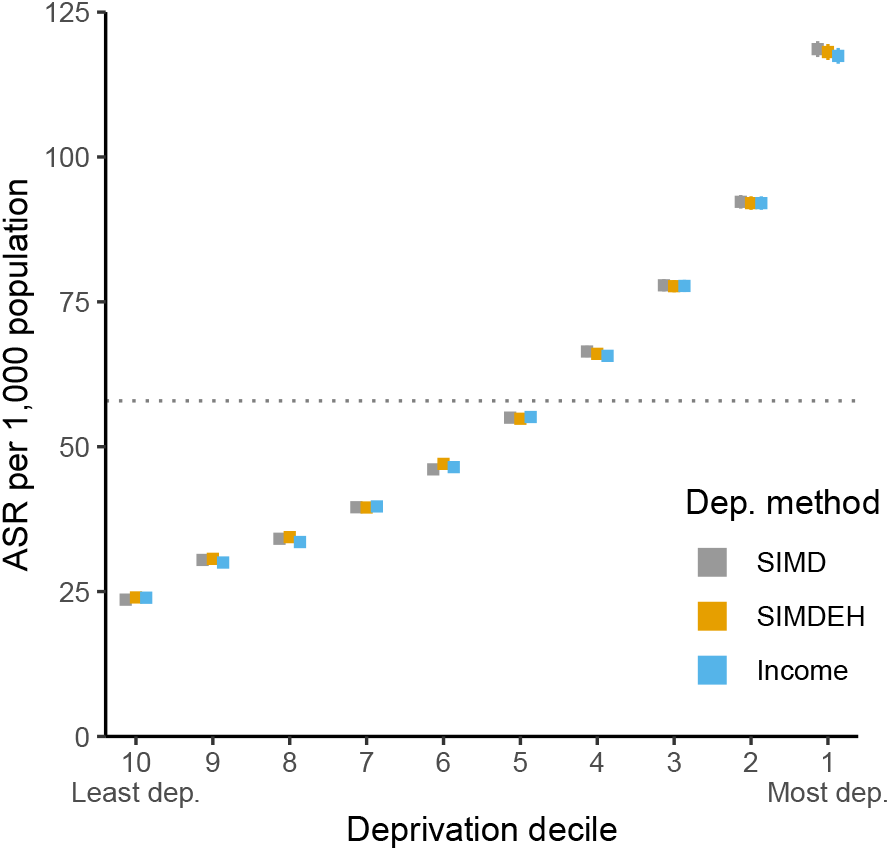
Age-standardised prevalence rate of females self-reporting bad or very bad health per 1,000 population. The rate for the population as whole is represented with a black dashed horizontal line (57.9 per 1,000 population; 95% CI: 57.6, 58.2). Note: The size of printed points is larger than the confidence intervals for most deciles.

No statistically significant difference in SII between SIMD and either of the alternative deprivation methods was found in any combination of age group, outcome, and sex. Absolute inequalities in health outcomes exhibited a linear increase from early adulthood to age 55–59 years for females (Figure 2), where it plateaued for self-reported bad health, and began to decrease rapidly for LLTHC with increasing age. The pattern in men was broadly similar except peak values of inequalities were found in those aged 60–64 years. The scale of absolute inequalities for these outcomes were similar between males and females in all age groups and subgroups. No statistically significant difference in RII between SIMD and either of the alternative deprivation methods was found in any combination of age group, outcome, and sex. Relative inequalities for health outcomes were moderately high in early childhood and lowest between age 10–30 years. Peak inequalities occurred in, or close to, the 40–44 year age group before decreasing into older adulthood. Relative inequalities were slightly larger in magnitude in middle-aged males than females but were otherwise similarly patterned. Figure 3 is presented as an example of this patterning with age and shows the relative inequality in rates of females living with a LLTHC. Relative inequalities in self-reported bad health were substantially larger than for rates of individuals living with LLTHC, with values of RII for bad health ranging between 1 and 2.5. These values indicate the rates of bad health in the most deprived data zones are about 50–125% higher than the least deprived across age groups (Authors, 2019c, p.36). Focusing on the second objective of this study, Figures 2 and 3 clearly demonstrate the high degree of overlap between all deprivation methods.

**Figure 2:**
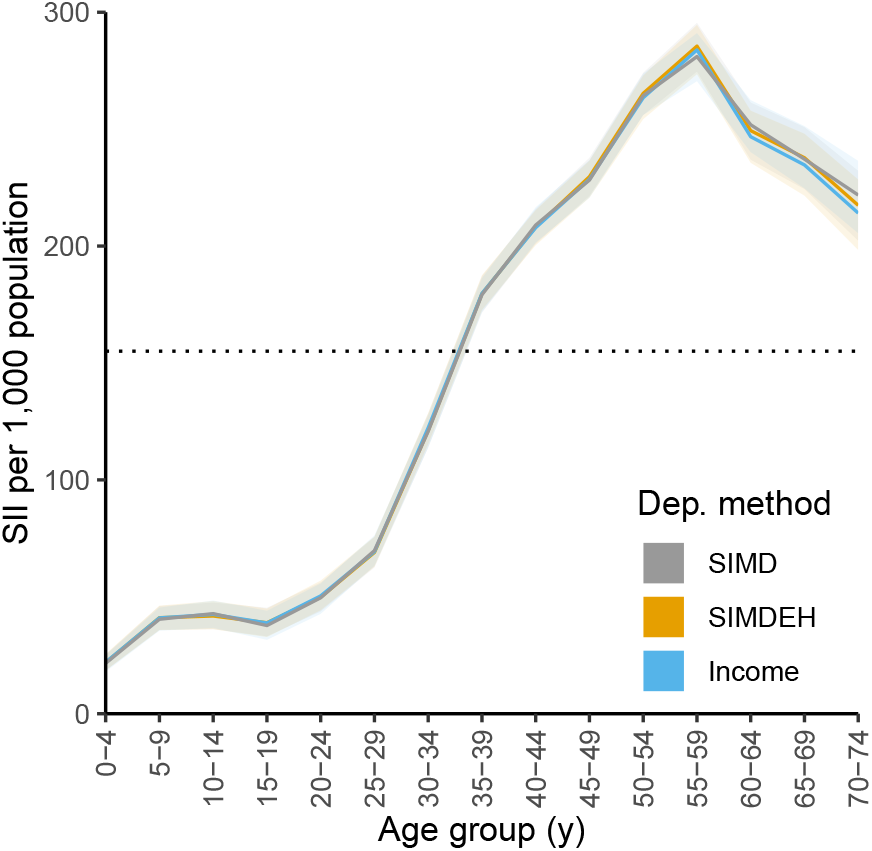
Age-standardised prevalence rate of females living with a limiting long-term health conditions (LLTHC) by age group. Point estimates and 95% confidence bands of absolute inequalities as indicated by the Slope Index of Inequality (SII). The SII for the population as a whole is represented with a black dashed horizontal line, estimated using overall Scottish Index of Multiple Deprivation decile midpoint values (155.0 per 1,000 population; 95% CI: 152.6, 157.3).

**Figure 3:**
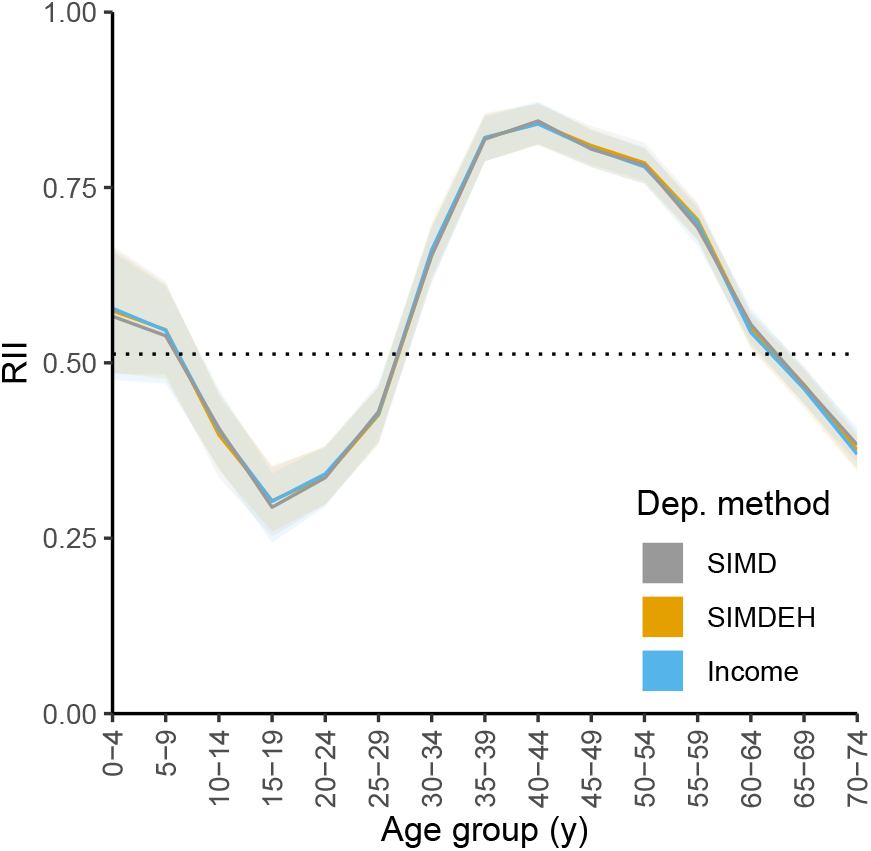
Age-standardised prevalence rate of females living with a limiting long-term health conditions (LLTHC) by age group. Point estimates and 95% confidence bands of relative inequalities as indicated by the Relative Index of Inequality (RII). The RII for the population as a whole is represented with a black dashed horizontal line, estimated using overall Scottish Index of Multiple Deprivation decile midpoint values (0.51; 95% CI: 0.50, 0.53).

### 3.3. Mortality

Descriptive statistics for mortality outcomes are reported in Table 2. Standardised mortality rates for males were significantly higher than for females. Standardised mortality rates for the overall population for both males and females decreased significantly between 2011 and 2018, but the absolute difference between males and females remained consistent.

Absolute inequalities in mortality increased exponentially with age, driven by the expected increase in mortality rate with age (see Supplement File Figure S5). Relative inequalities, however, peaked at ages 30–49 and decreased slowly with age for both males and females (see Supplement File Figure S6). Absolute inequalities in females increased significantly from 2011 to 2018 but no such difference was found in males (Table 3). Relative inequality significantly increased in both females and males between time periods. Premature mortality exhibited larger greater relative inequalities than all-age mortality (results not shown). Both absolute and relative inequalities in individuals aged 30–54 years demonstrated an increase between 2011 and 2018, although differences in relative inequalities were not statistically significant between time periods for any single five-year age group. These results once again highlight that the magnitude of differences in calculated values between deprivation methods are insubstantial in comparison to differences between age groups or time periods.

### 3.4. Auxiliary results

No substantial differences in the general patterning of health nor mortality outcomes were noted in the wider set of unreported results which used different numbers of quantiles (quintiles and vigintiles) or other isolated domains of the SIMD. The exception to this is the access deprivation domain. Increased access-related deprivation was associated with improved health and mortality outcomes. An example plot is shown in Supplement File Figure S7 which presents standardised rates for self-rated bad health for all deprivation methods by decile. This reversal of trend is due to access-related deprivation domain scores being based on indicators such as the availability of high-speed internet and travel times to a range of key education and healthcare services. Materially deprived areas tend to cluster in urban centres which have a higher density of such services. This leads to a moderate negative correlation between deprivation rank in the access domain and all other constituent domains of the SIMD, as well as composite deprivation rank (Supplement File Tables S2 and S3).

## 4. Discussion

There is ongoing concern that the inclusion of health-related indicators in composite measures of multiple deprivation (CMMD) can lead to endogeneity bias in health inequalities analyses. This bias may ultimately lead to inappropriate allocation of healthcare resources and the unintentional perpetuation or worsening of health inequalities. Our work addressed this concern and found no substantial differences between the two commonly-used bias mitigation strategies (exclusion of the health domain and using the income domain in isolation). The results of the present study demonstrated that the risk of endogeneity bias in health inequalities research due to inclusion of health-related indicators in CMMDs is negligible.

### 4.1. Comparison of deprivation calculation methods

This study used population-level health and mortality data, and a CMMD in a country with significant health inequalities. By using these data in a selection of analysis methods frequently found in health inequalities research, we have demonstrated the magnitude of inequalities between socioeconomic strata outweigh differences between methods of calculating deprivation within a single stratum. This finding broadly persisted regardless of whether quintiles, deciles, or vigintiles were selected as the unit of quantile division.

The underlying reason for the lack of difference between mitigation strategies is the strong association between the health domain and overall deprivation ranks (*r*_*s*_ = .91 and .94 for SIMD2012 and SIMD2020v2 respectively; see Supplementary Tables S2 and S3 for all interdomain correlation values). There is therefore minimal impact on a data zone’s composite deprivation rank, and the quantile to which is it subsequently assigned, regardless if the health domain is included or not. The income domain is similarly highly correlated with overall deprivation (*r*_*s*_ = .97 for both iterations of the SIMD). Thus, using this domain in isolation does not change the quantile to which the majority of data zones are assigned. The majority of other domains are also strongly associated with the overall deprivation rank (excluding the access domain, as discussed above). This strong interdomain correlation is unsurprising since indicators included in CMMDs are selected specifically because they exhibit socioeconomic gradients. Researchers using CMMDs other than the SIMD, such as the Income and Employment Index (IEI) used in the Scottish Goverment’s Long-term Monitoring of Health Inequalities report (Scottish Government, 2022), can therefore quickly address any concerns about potential endogeneity bias by considering the strength of interdomain correlation in their chosen measure.

Our results extend those of Adams and White (2006) by demonstrating a lack of substantial difference between deprivation for more contemporary health data after excluding the health domain of our chosen CMMD. Our work also complements past research that has considered varying the weights applied to constituent domains (which have been the source of debate; Deas et al., 2003; Dibben et al., 2007; Watson et al., 2019). Schederecker et al. (2019) compared a range of methods for weighting domains of the German Index of Multiple Deprivation. Their study found that even substantial changes in domain weights produced little difference in the association of mortality rates with deprivation. Strömberg et al. (2021) constructed a CMMD for use in Sweden and found it as efficacious as a measure of deprivation as household income, although the former did outperform other typical single-item measures. Other research comparing different indices of deprivation indicates that selection of measure, or varying the size of geographic zones used, can have much more notable effects on results than implementing the mitigation strategies as we have in the present work (Adams et al., 2005; Berkowitz et al., 2015; Lian et al., 2016). However, a comparison of seven CMMDs by Bryere et al. (2017) demonstrated that most were comparable in the degree to which they agreed with single-item measures (e.g., highest educational attainment or household income). Another recent study in New Zealand also demonstrated that different CMMDs adequately capture deprivation even when underpinned by different theoretical positions and constituent indicators (Crampton et al., 2020). Together, these results indicate area-level deprivation can be suitably quantified in many ways and the results of health inequalities analyses are robust, regardless of differences in deprivation measurement methodologies.

### 4.2. Health outcomes

Health inequalities due to deprivation were substantial and pervasive across health outcomes in this study, with increased deprivation being associated with poorer health. Differences between strata (deprivation deciles and age groups) were far larger in magnitude than differences between deprivation methods within the same stratum. This reinforces that the risk of endogeneity bias due to the inclusion of health domains in CMMD is minimal.

The proportion of people living with self-rated bad health increased both with age and with deprivation. However, only 15% of people aged 75 or above consider themselves to be in bad or very bad health despite nearly 75% of both females and males having at least one limiting long-term health condition in this age group. The crude proportion of individuals reporting self-rated fair, bad, or very bad health (calculated but not reported here) was substantially lower than the rates reported by Adams et al. (2005). However, that study was limited to a northern English city that was not representative of the national population in terms of deprivation. Standardised rates in the present study are comparable to previous work which used 2011 Scottish Census health data (Authors, 2019b).

### 4.3. Mortality

Our results demonstrated a lack of substantial difference in mortality outcomes between deprivation methods, an outcome which has not been previously explored in the literature in the context of endogeneity bias in CMMDs. Differences between strata of deprivation or age were, again, larger in magnitude than differences between deprivation methods within the same stratum.

Age-standardised mortality rates substantially worsened with increasing deprivation for both females and males, for both time periods of mortality analysis. Absolute inequalities in mortality rates measured using the Slope Index of Inequality increased with age. A significant increase in absolute inequalities was observed between 2011 and 2018 in both middle-aged females and males. Relative inequalities measured using the Relative Index of Inequality peaked in middle age for both females and males but were generally higher in males, in keeping with a previous study of mortality in Scotland (Ralston et al., 2014). There was a significant decrease in age-standardised mortality rates for both females and males between the two time periods, as has been found in other parts of the UK (Murphy, 2021). Overall relative inequalities in 2018 increased significantly from 2011 levels, particularly for females. However, the difference was not statistically significant between time periods for either sex for any individual five year age group with the exception of males aged 45–49 years (see Supplemental File Figure S6). The increase in mortality for middle-aged men may be explained in part by the increase in deaths due to suicide, alcohol, and drug use (Authors, 2020).

### 4.4. Strengths and limitations

A strength of our work is the wide range of analyses carried out for various subgroups (age groups, sex, socioeconomic strata) for multiple health and mortality outcomes using population-level data. By exploring a selection of age subgroups and different numbers of quantiles using common analytical methods we have demonstrated the robustness of health inequalities research to commonly-used bias mitigation strategies used in deprivation measurement methodology. Additionally, employing a widely-used CMMD in a pragmatic manner grounds the present work in realworld applications of CMMDs. This pragmatism means the present work is of value to health inequalities researchers, healthcare resourcing, and governmental organisations.

The present work is limited by its use of a single CMMD. The weights applied to each of the constituent domains and indicators of the SIMD are potentially idiosyncratic to Scotland and may limit generalisability. Future work using alternative CMMDs with different weight distributions, or carrying out a sensitivity analysis, can address this limitation. Further, self-rated health outcomes are subjective and at least partly constructed by an individual comparing themselves to others in close proximity. As such, absolute differences in health-related quality of life across the population may be inaccurately reflected by self-rated health outcomes. This could lead to an underestimate of the degree of deprivation-related health inequalities. Objective measures of health conditions or outcomes (e.g., disability-adjusted life years) would address this limitation. Census data are also not collected for individuals living in communal dwellings.

### 4.5. Conclusion

Our work indicates the risk of endogeneity bias in relation to the use of CMMDs in health inequalities research is negligible. Moreover, it suggests that current practices surrounding the use of the SIMD and similar CMMDs appropriately capture health inequalities due to differences in deprivation at the area level. Mitigation strategies to avoid potential endogeneity bias may be theoretically appropriate and prevent health appearing simultaneously in outcome and predictor variables, but these strategies appear to have minimal practical impact. Researchers must continue to consider which method of measuring deprivation is the most appropriate to the problem at hand and if mitigation strategies are worthwhile. However, there is unlikely to be any substantial difference in results when using common variations of a particular CMMD. Indeed, the selection of measure (e.g, Scottish Index of Multiple Deprivation v. single-item indicators) will likely have far more impact on results. Even then, the degree of health inequalities between the most- and least-deprived areas will in many cases be far larger than variation between deprivation measures. Researchers should be reassured by the present work that the selection of deprivation measure should be of relatively minor concern in their study design. Focus must remain on addressing preventable health inequalities.

## Data Availability

See manuscript for data statement.

## CRediT authorship contribution statement

**D R R Bradford:** Conceptualization, methodology, software, formal analysis, investigation, resources, writing – original draft, writing – review and editing, visualization, project administration. **M Allik:** Writing – review and editing, supervision. **A D McMahon:** Writing – review and editing, supervision. **D Brown:** Resources, writing – review and editing, supervision.

## Funding

This research was funded by the Medical Research Council and Scottish Chief Scientist’s Office as part of an PhD studentship awarded to DRRB [grant MC_ST_00022]. This research was funded by the Medical Research Council and Scottish Chief Scientist’s Office as part of an PhD studentship awarded to DRRB [grant MC_ST_00022]. MA and DB were the Medical Research Council [grant MC_UU_00022/2] and the Scottish Government Chief Scientist’s Office [grant SPHSU17]. MA was also supported by the Economic and Social Research Council [grant ES/T000120/1].

## Acknowledgements

DRRB would like to thank Elizabeth Fraser of the Scottish Government Communities Analysis Division for useful discussions of the methods used to calculate Scottish Index of Multiple Deprivation scores and ranks. DRRB would also like to thank Andreas Hoehn and Daniel Kopasker of the MRC/CSO Social and Public Health Sciences Unit, University of Glasgow, for their critical review of this work.

---

**Table S1.** Indicators comprising the health domain of the 2012 and 2020v2 iterations of the Scottish Index of Multiple Deprivation and their relative weights. See the technical notes provided by the Scottish Government for further detail (Scottish Government, 2012, 2020).

| *Indicator* | *2012* | *2020v2* |
| --- | --- | --- |
| *Standardised mortality ratio* | .09 | .06 |
| *Hospital stays related to alcohol use: standardised ratio* | .15 | .08 |
| *Hospital stays related to drug use: standardised ratio* | .07 | .07 |
| *Comparative illness factor: standardised ratio* | .14 | .46 |
| *Emergency stays in hospital: standardised ratio* | .47 | .19 |
| *Proportion of population being prescribed drugs for anxiety, depression or psychosis* | .05 | .13 |
| *Proportion of live singleton births of low birth weight* | .02 | .01 |

**Table S2.** Spearman’s rank interdomain correlation coefficients for 2012 iteration of the Scottish Index of Multiple Deprivation (SIMD).

|  | *SIMD* | *SIMDEH* | *Income* | *Employment* | *Health* | *Education* | *Crime* | *Access* | *Housing* |
| --- | --- | --- | --- | --- | --- | --- | --- | --- | --- |
| *SIMD* | 1.00 | 1.00 | 0.97 | 0.97 | 0.91 | 0.92 | 0.72 | -0.23 | 0.70 |
| *SIMDEH* | 1.00 | 1.00 | 0.96 | 0.96 | 0.87 | 0.92 | 0.71 | -0.21 | 0.70 |
| *Income* | 0.97 | 0.96 | 1.00 | 0.96 | 0.89 | 0.89 | 0.71 | -0.34 | 0.68 |
| *Employment* | 0.97 | 0.96 | 0.96 | 1.00 | 0.89 | 0.88 | 0.71 | -0.33 | 0.65 |
| *Health* | 0.91 | 0.87 | 0.89 | 0.89 | 1.00 | 0.81 | 0.70 | -0.35 | 0.63 |
| *Education* | 0.92 | 0.92 | 0.89 | 0.88 | 0.81 | 1.00 | 0.67 | -0.27 | 0.69 |
| *Crime* | 0.72 | 0.71 | 0.71 | 0.71 | 0.70 | 0.67 | 1.00 | -0.50 | 0.59 |
| *Access* | -0.23 | -0.21 | -0.34 | -0.33 | -0.35 | -0.27 | -0.50 | 1.00 | -0.38 |
| *Housing* | 0.70 | 0.70 | 0.68 | 0.65 | 0.63 | 0.69 | 0.59 | -0.38 | 1.00 |

**Table S3.** Spearman’s rank interdomain correlation coefficients for the 2020v2 iteration of Scottish Index of Multiple Deprivation (SIMD).

|  | *SIMD* | *SIMDEH* | *Income* | *Employment* | *Health* | *Education* | *Crime* | *Access* | *Housing* |
| --- | --- | --- | --- | --- | --- | --- | --- | --- | --- |
| *SIMD* | 1.00 | 1.00 | 0.97 | 0.97 | 0.94 | 0.91 | 0.66 | -0.23 | 0.69 |
| *SIMDEH* | 1.00 | 1.00 | 0.97 | 0.96 | 0.92 | 0.91 | 0.66 | -0.20 | 0.69 |
| *Income* | 0.97 | 0.97 | 1.00 | 0.96 | 0.94 | 0.88 | 0.64 | -0.34 | 0.70 |
| *Employment* | 0.97 | 0.96 | 0.96 | 1.00 | 0.94 | 0.86 | 0.64 | -0.34 | 0.66 |
| *Health* | 0.94 | 0.92 | 0.94 | 0.94 | 1.00 | 0.86 | 0.62 | -0.35 | 0.65 |
| *Education* | 0.91 | 0.91 | 0.88 | 0.86 | 0.86 | 1.00 | 0.60 | -0.27 | 0.68 |
| *Crime* | 0.66 | 0.66 | 0.64 | 0.64 | 0.62 | 0.60 | 1.00 | -0.44 | 0.59 |
| *Access* | -0.23 | -0.20 | -0.34 | -0.34 | -0.35 | -0.27 | -0.44 | 1.00 | -0.43 |
| *Housing* | 0.69 | 0.69 | 0.70 | 0.66 | 0.65 | 0.68 | 0.59 | -0.43 | 1.00 |

**Table S4.** Distribution of decile shifts experienced by data zones for alternative deprivation methods. *N*_*D*_ = number of deciles shifted relative to each data zone’s original assigned decile using the overall Scottish Index of Multiple Deprivation (SIMD).

| $N_D$ | 2012 | | 2020v2 | |
| --- | --- | --- | --- | --- |
|  | *SIMDEH* | *Income* | *SIMDEH* | *Income* |
| -3 | 0 | 5 | 0 | 3 |
| -2 | 0 | 194 | 0 | 152 |
| -1 | 603 | 1,417 | 473 | 1,388 |
| 0 | 5,373 | 3,451 | 6,036 | 3,971 |
| 1 | 528 | 1,204 | 467 | 1,228 |
| 2 | 1 | 214 | 0 | 217 |
| 3 | 0 | 15 | 0 | 14 |
| 4 | 0 | 5 | 0 | 3 |
| *Sum* | 6,505 | 6,505 | 6,976 | 6,976 |

**Figure S1:**
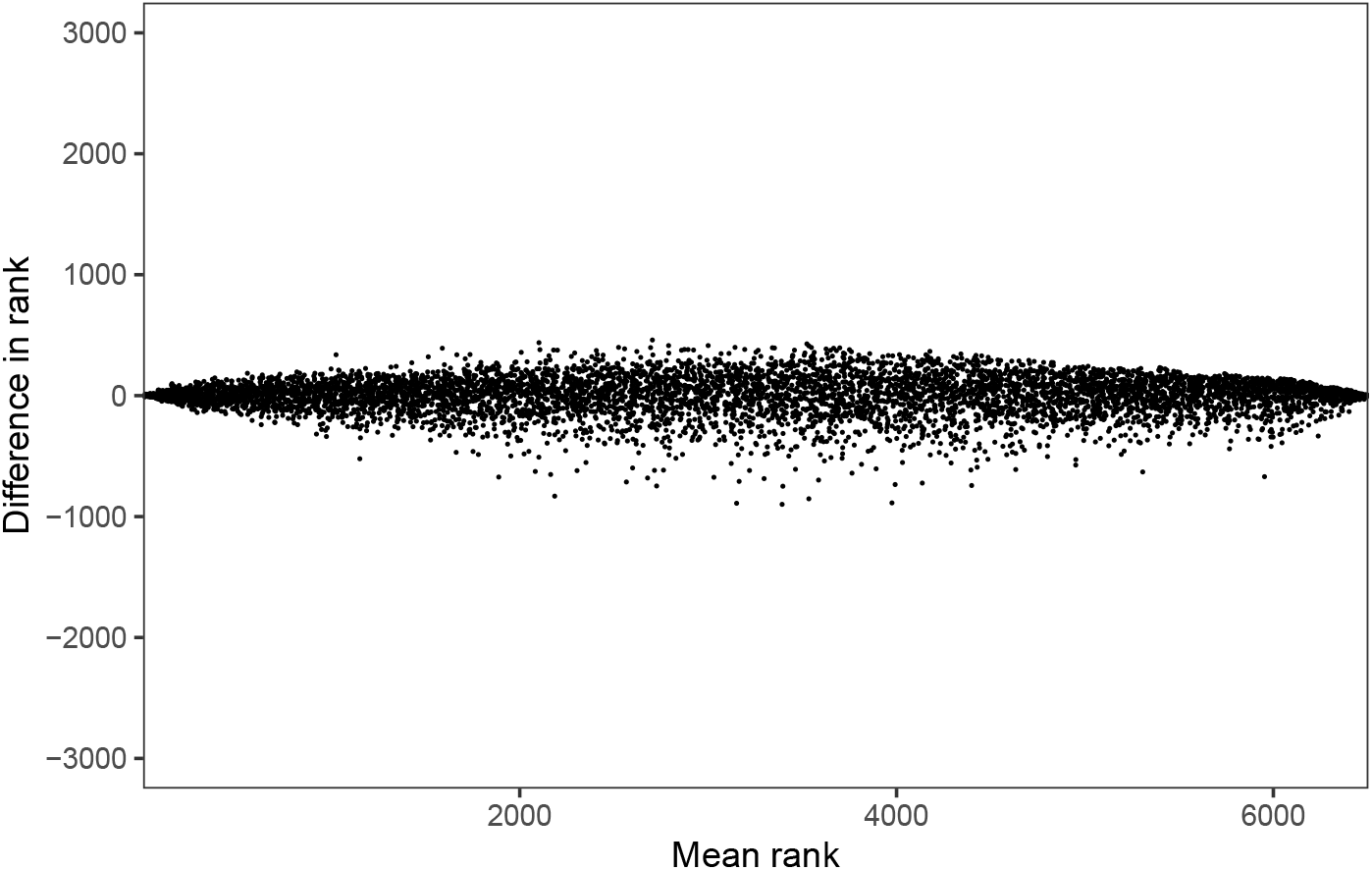
Bland–Altman plot showing pairwise difference in ranks and pairwise mean rank of the overall and health-excluded versions of the 2012 iteration of the Scottish Index of Multiple Deprivation (SIMD) for all data zones.

**Figure S2:**
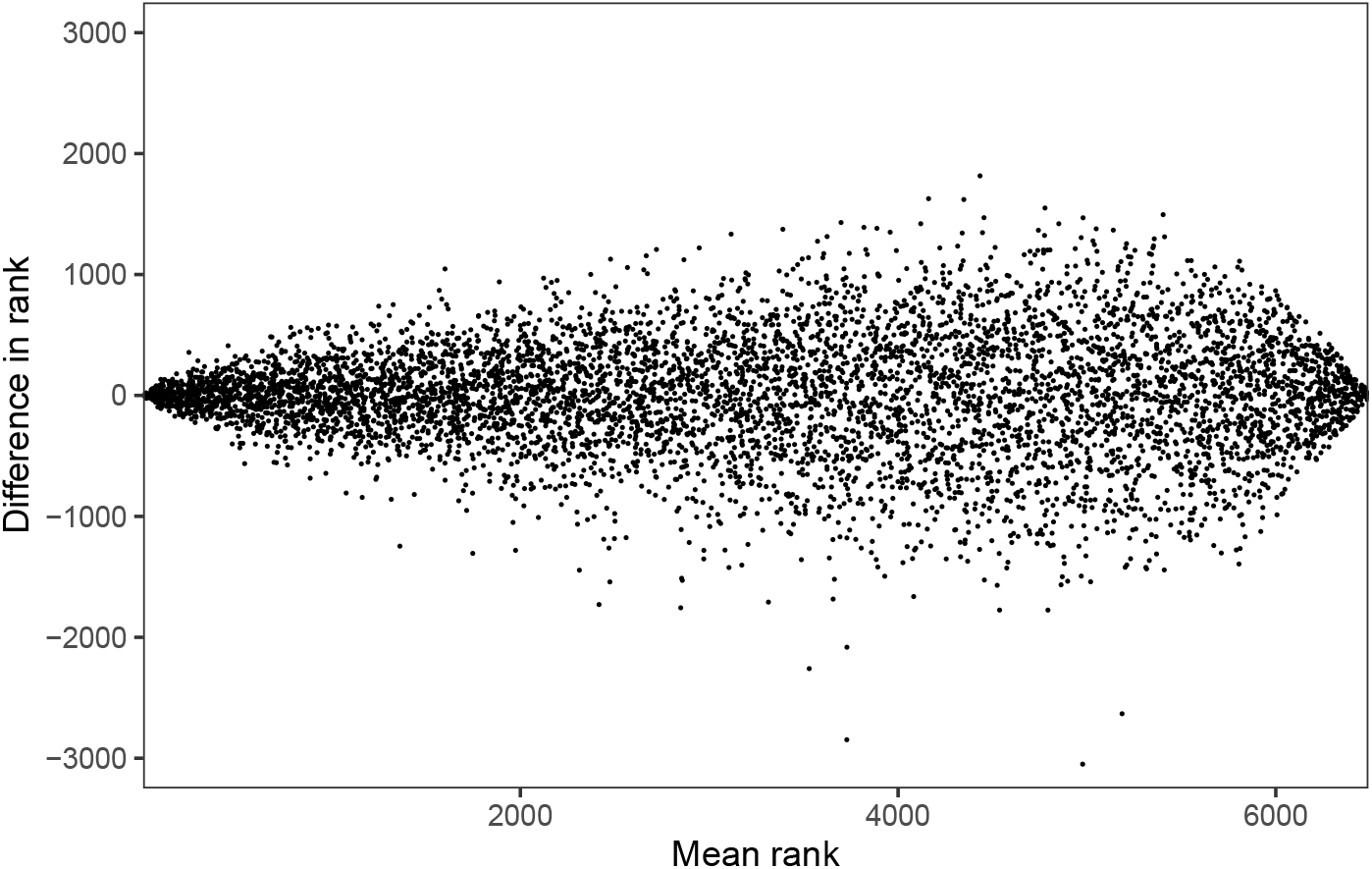
Bland–Altman plot showing pairwise difference in ranks and pairwise mean rank of the overall and income domains of the 2012 iteration of the Scottish Index of Multiple Deprivation (SIMD) for all data zones.

**Figure S3:**
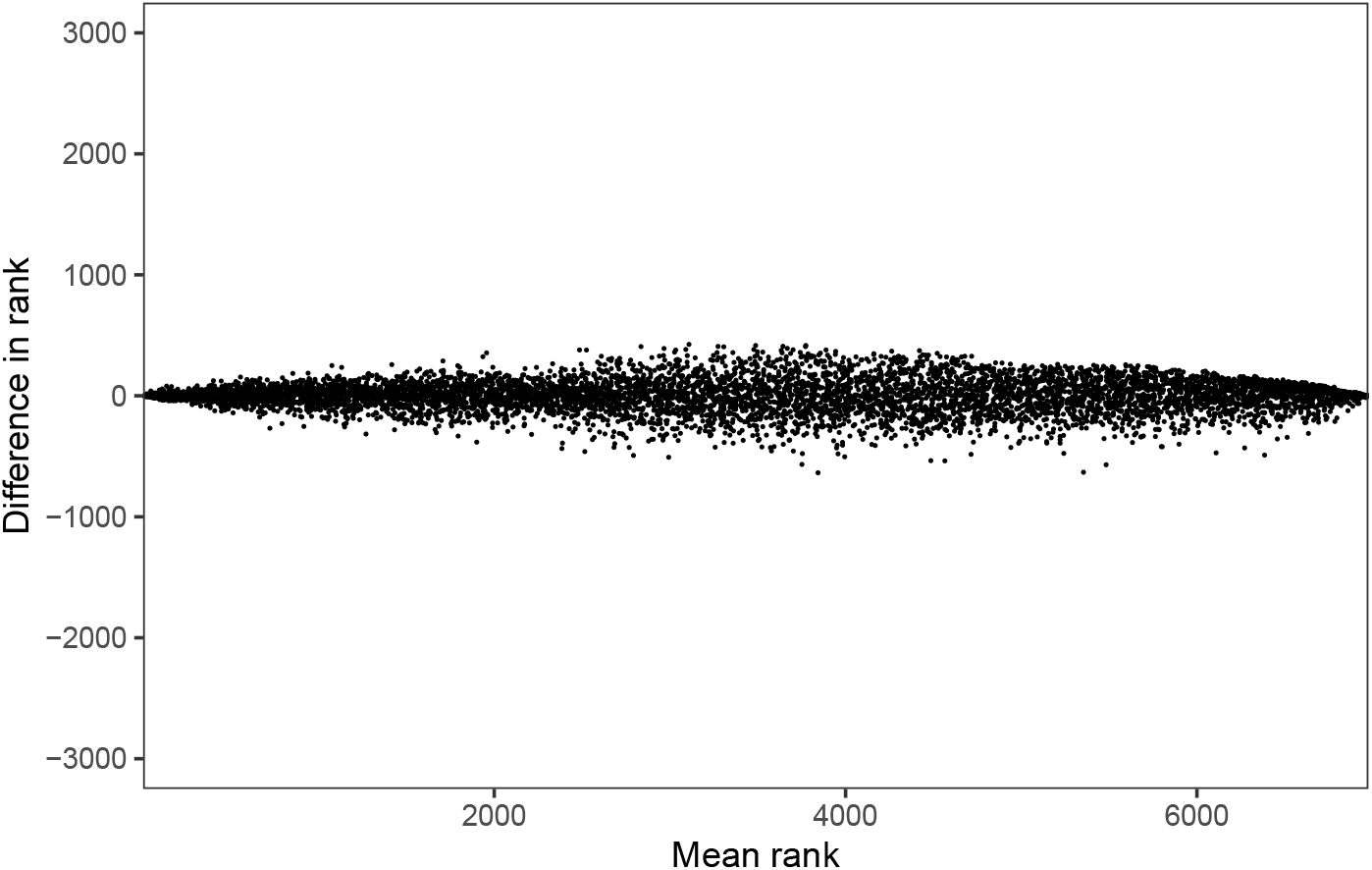
Bland–Altman plot showing pairwise difference in ranks and pairwise mean rank of the overall and health-excluded versions of the 2020v2 iteration of the Scottish Index of Multiple Deprivation (SIMD) for all data zones.

**Figure S4.**
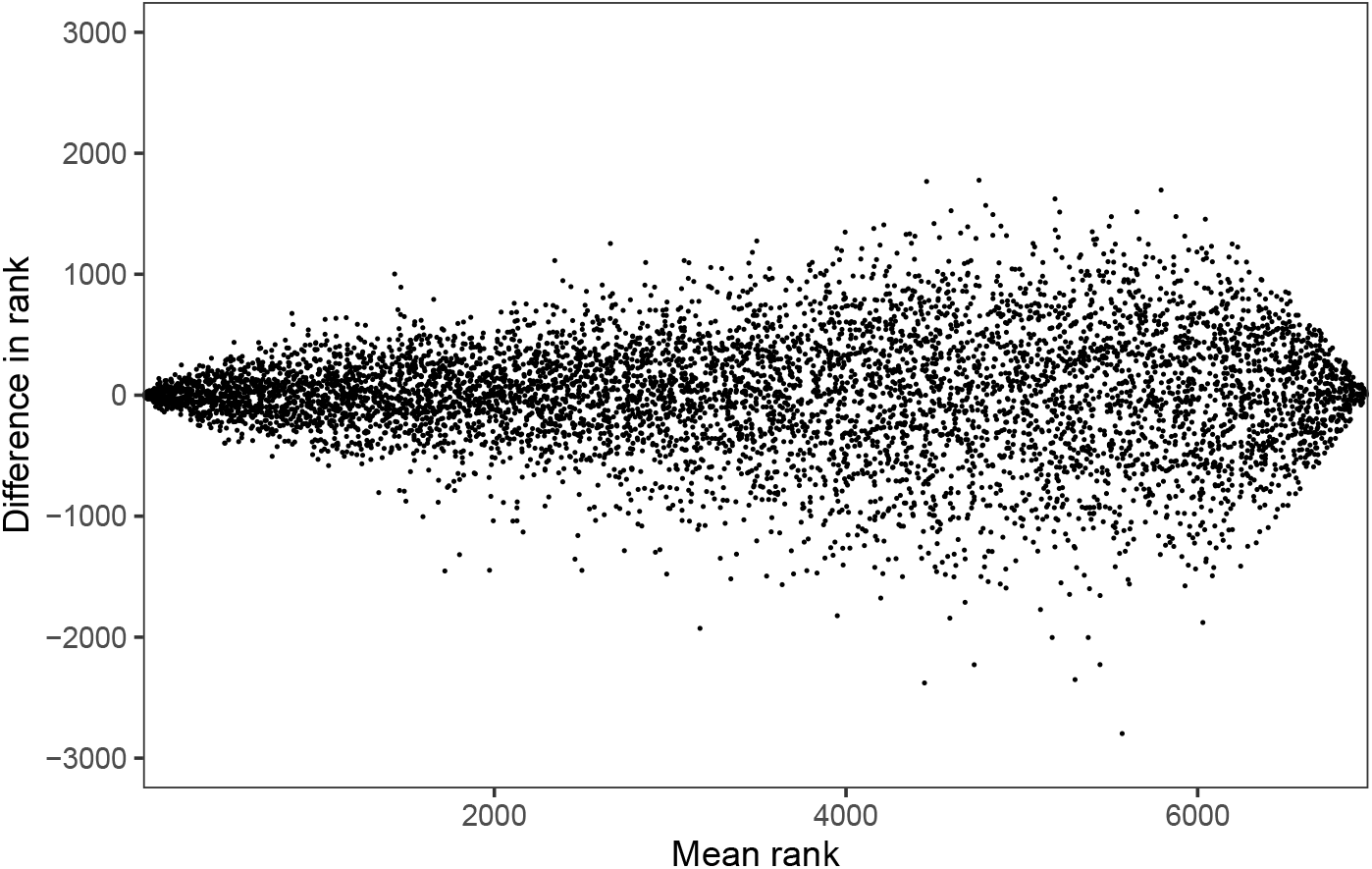
Bland–Altman plot showing pairwise difference in ranks and pairwise mean rank of the overall and income domains of the 2020v2 iteration of the Scottish Index of Multiple Deprivation (SIMD) for all data zones.

**Figure S5:**
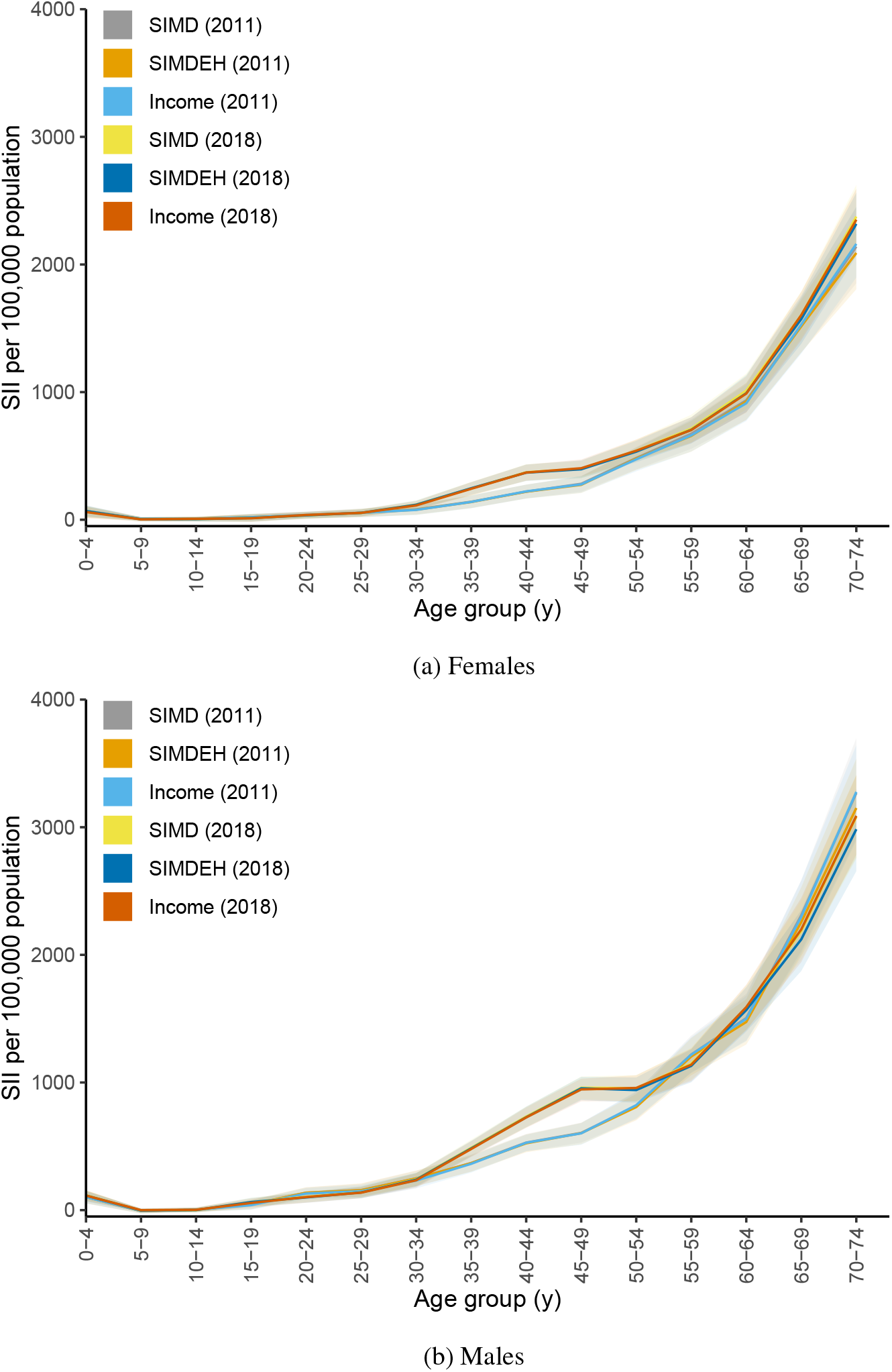
Point estimates and 95% confidence bands of absolute inequalities (as indicated by the Slope Index of Inequality; SII) in the standardised mortality rates in 2011 and 2018 by five-year age groups.

**Figure S6.**
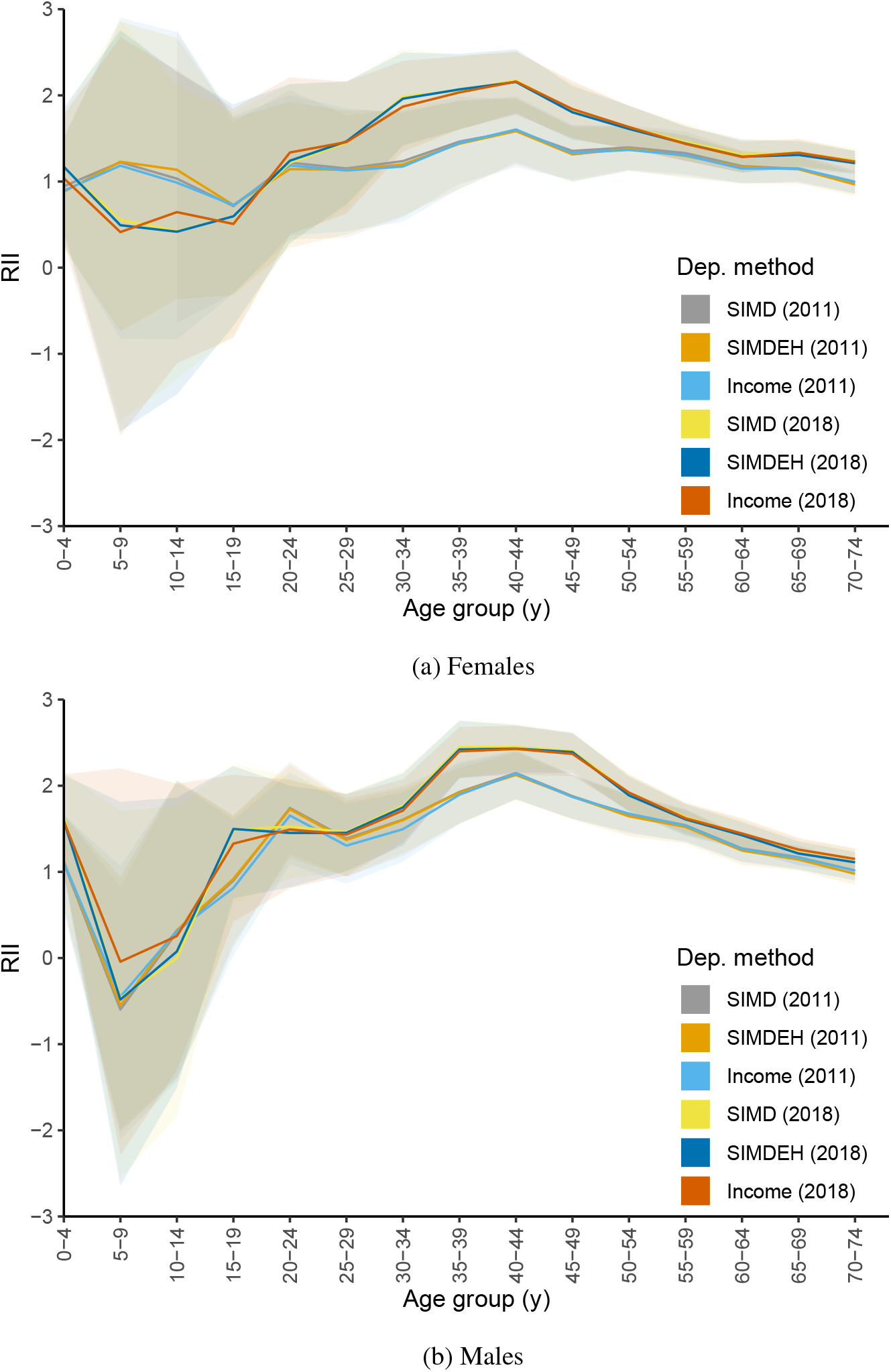
Point estimates and 95% confidence bands of relative inequalities (as indicated by the Relative Index of Inequality; RII) in the standardised mortality rates in 2011 and 2018 by five-year age groups.

**Figure S7:**
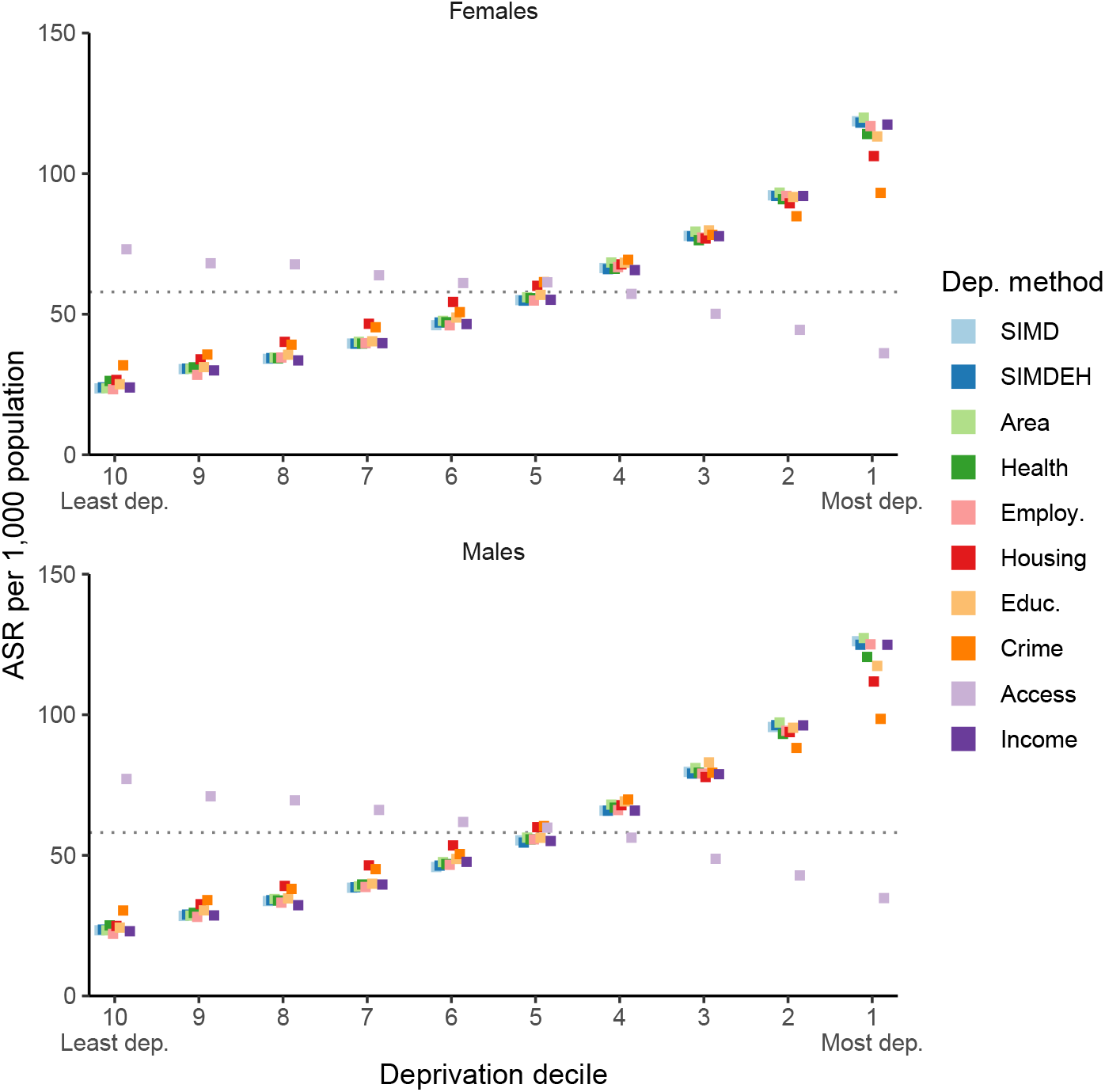
Age-standardised prevalence rate of individuals self-reporting bad or very bad health. The rate for the population as a whole is represented by the broken horizontal line; females = 57.9 (95% CI: 57.6, 58.21) per 1,000 population; malesm= 58.1 (95% CI: 57.8, 58.4) per 1,000 population.

## Notes

### Competing Interest Statement

The authors have declared no competing interest.

### Funding Statement

See manuscript for funding statement.

## References

Adams, J., Ryan, V., White, M., 2005. How accurate are Townsend Deprivation Scores as predictors of self-reported health? a comparison with individual level data. J. Public Health 27, 101–106. doi:10.1093/pubmed/fdh193.

Adams, J., White, M., 2006. Removing the health domain from the index of multiple deprivation 2004—effect on measured inequalities in census measure of health. J. Public Health 28, 379–383. doi:10.1093/pubmed/fdl061.

Agardh, E., Allebeck, P., Hallqvist, J., Moradi, T., Sidorchuk, A., 2011. Type 2 diabetes incidence and socio-economic position: a systematic review and meta-analysis. Int. J. Epidemiol. 40, 804–818. doi:10.1093/ije/dyr029.

Archie, J.P., 1981. Mathematic coupling of data. Ann. Surg. 193, 296–303. doi:10.1097/00000658-198103000-00008.

Australian Bureau of Statistics, 2018. Socio-Economic Indexes for Areas (SEIFA): Technical Paper. URL: https://www.ausstats.abs.gov.au/ausstats/subscriber.nsf/0/756EE3DBEFA869EFCA258259000BA746/$File/SEIFA2016TechnicalPaper.pdf.

Authors,.

Authors, 2009. Journal [details removed for peer review].

Authors, 2016. Journal [details removed for peer review].

Authors, 2018. Journal [details removed for peer review].

Authors, 2019a. Journal [details removed for peer review].

Authors, 2019b. Journal [details removed for peer review].

Authors, 2019c. Chapter [details removed for peer review].

Authors, 2020. Journal [details removed for peer review].

Authors, 2021. Journal [details removed for peer review].

Authors, 2022. Journal [details removed for peer review].

Baczyk, M., Schenk, K., McLaughlin, D., McGuire, A., Gadsden, S., 2016. Place-based approaches to joint planning, resourcing and delivery - An overview of current practice in Scotland. URL: http://www.improvementservice.org.uk/documents/research/place-based-approaches-report.pdf.

Berkowitz, S.A., Traore, C.Y., Singer, D.E., Atlas, S.J., 2015. Evaluating Area-Based Socioeconomic Status Indicators for Monitoring Disparities within Health Care Systems: Results from a Primary Care Network. Health Serv. Res. 50, 398–417. URL: https://onlinelibrary.wiley.com/doi/10.1111/1475-6773.12229, doi:10.1111/1475-6773.12229.

Braveman, P.A., Cubbin, C., Egerter, S., Chideya, S., Marchi, K.S., Metzler, M., Posner, S., 2005. Socioeconomic Status in Health Research. JAMA 294, 2879. doi:10.1001/jama.294.22.2879.

Bryere, J., Pornet, C., Copin, N., Launay, L., Gusto, G., Grosclaude, P., Delpierre, C., Lang, T., Lantieri, O., Dejardin, O., Launoy, G., 2017. Assessment of the ecological bias of seven aggregate social deprivation indices. BMC Public Health 17, 86. doi:10.1186/s12889-016-4007-8.

Cohen, J., 1968. Weighted kappa: Nominal scale agreement provision for scaled disagreement or partial credit. Psychol. Bull. 70, 213–220. doi:10.1037/h0026256.

Covvey, J.R., Johnson, B.F., Elliott, V., Malcolm, W., Mullen, A.B., 2014. An association between socioeconomic deprivation and primary care antibiotic prescribing in Scotland. J. Antimicrob. Chemother. 69, 835–841. doi:10.1093/jac/dkt439.

Crampton, P., Salmond, C.E., Atkinson, J., 2020. A comparison of the NZDep and New Zealand IMD indexes of socioeconomic deprivation. Kōtuitui New Zeal. J. Soc. Sci. Online 15, 154–169. doi:10.1080/1177083X.2019.1676798.

Deas, I., Robson, B., Wong, C., Bradford, M., 2003. Measuring neighbourhood deprivation: A critique of the Index of Multiple Deprivation. Environ. Plan. C Gov. Policy 21, 883–903. doi:10.1068/c0240.

Dibben, C., Atherton, I., Cox, M., Watson, V., Ryan, M., Sutton, M., 2007. Investigating the impact of changing the weights that underpin the Index of Multiple Deprivation 2004. URL: http://webarchive.nationalarchives.gov.uk/20100410180038/http://www.communities.gov.uk/publications/communities/investigatingimpact.

Eurostat, 2013. Revision of the European Standard Population. URL: https://ec.europa.eu/eurostat/web/products-manuals-and-guidelines/-/ks-ra-13-028.

Exeter, D.J., Zhao, J., Crengle, S., Lee, A., Browne, M., 2017. The New Zealand Indices of Multiple Deprivation (IMD): A new suite of indicators for social and health research in Aotearoa, New Zealand. PLoS One 12, e0181260. doi:10.1371/journal.pone.0181260.

Fenton, L., Wyper, G.M., McCartney, G., Minton, J., 2019. Socioeconomic inequality in recent adverse all-cause mortality trends in Scotland. J. Epidemiol. Community Health 73, 971–974. doi:10.1136/jech-2019-212300.

Frølich, A., Ghith, N., Schiøtz, M., Jacobsen, R., Stockmarr, A., 2019. Multimorbidity, healthcare utilization and socioeconomic status: A register-based study in Denmark. PLoS One 14, e0214183. doi:10.1371/journal.pone.0214183.

Georgina, A., 2020. PHEindicatormethods: Common Public Health Statistics and their Confidence Intervals. URL: https://CRAN.R-project.org/package=PHEindicatormethods. r package version 1.3.2.

Harrell Jr, F.E., 2021. Hmisc: Harrell Miscellaneous. URL: https://CRAN.R-project.org/package=Hmisc. r package version 4.6-0.

Henery, P.M., Dundas, R., Katikireddi, S.V., Leyland, A., Wood, R., Pearce, A., 2021. Social inequalities and hospital admission for unintentional injury in young children in Scotland: A nationwide linked cohort study. Lancet Reg. Heal. - Eur. 6, 100117. doi:10.1016/j.lanepe.2021.100117.

Hill, A.D., Johnson, S.G., Greco, L.M., O’Boyle, E.H., Walter, S.L., 2021. Endogeneity: A review and agenda for the methodology-practice divide affecting micro and macro research. J. Manage. 47, 105–143. doi:10.1177/0149206320960533.

Jordan, H., 2004. The Index of Multiple Deprivation 2000 and accessibility effects on health. J. Epidemiol. Community Heal. 58, 250–257. doi:10.1136/jech.2003.013011.

Knies, G., Kumari, M., 2022. Multimorbidity is associated with the income, education, employment and health domains of area-level deprivation in adult residents in the UK. Sci. Rep. 12, 7280. doi:10.1038/s41598-022-11310-9.

Lian, M., Struthers, J., Liu, Y., 2016. Statistical Assessment of Neighborhood Socioeconomic Deprivation Environment in Spatial Epidemiologic Studies. Open J. Stat. 6, 436–442. doi:10.4236/ojs.2016.63039.

Liberatos, P., Link, B.G., Kelsey, J.L., 1988. The measurement of social class in epidemiology. Epidemiol. Rev. 10, 87–121. doi:10.1093/oxfordjournals.epirev.a036030.

Mackenbach, J.P., Stirbu, I., Roskam, A.J.R., Schaap, M.M., Menvielle, G., Leinsalu, M., Kunst, A.E., 2008. Socioeconomic inequalities in health in 22 European countries. N. Engl. J. Med. 358, 2468–2481. doi:10.1056/NEJMsa0707519.

Maier, W., Holle, R., Hunger, M., Peters, A., Meisinger, C., Greiser, K.H., Kluttig, A., Völzke, H., Schipf, S., Moebus, S., Bokhof, B., Berger, K., Mueller, G., Rathmann, W., Tamayo, T., Mielck, A., 2013. The impact of regional deprivation and individual socio-economic status on the prevalence of Type 2 diabetes in Germany. A pooled analysis of five population-based studies. Diabet. Med. 30, e78–e86. doi:10.1111/dme.12062.

McCartney, G., Walsh, D., Whyte, B., Collins, C., 2012. Has Scotland always been the ‘sick man’ of Europe? An observational study from 1855 to 2006. Eur. J. Public Health 22, 756–760. doi:10.1093/eurpub/ckr136.

McLennan, D., Noble, S., Noble, M., Plunkett, E., Wright, G., Gutacker, N., 2019. The English Indices of Deprivation 2019: Technical Report. URL: https://assets.publishing.service.gov.uk/government/uploads/system/uploads/attachment_data/file/833951/IoD2019_Technical_Report.pdf.

Murphy, M., 2021. Recent mortality in Britain: a review of trends and explanations. Age and Ageing 50, 676–683. doi:10.1093/ageing/afab016.

National Records of Scotland,. Commissioned tables. URL: https://www.scotlandscensus.gov.uk/census-results/download-data/commissioned-tables/.

National Records of Scotland, 2022. Population Estimates by Scottish Index of Multiple Deprivation (SIMD). URL: https://www.nrscotland.gov.uk/statistics-and-data/statistics/statistics-by-theme/population/population-estimates/2011-based-special-area-population-estimates/population-estimates-by-simd-2016.

NHS England and Improvement, 2021. Technical Guide to Allocation Formulae and Convergence. URL: https://www.england.nhs.uk/wp-content/uploads/2022/04/technical-guide-to-integrated-care-board-allocations-22-23-to-24-25.pdf.

Northern Ireland Statistics and Research Agency, 2017. Northern Ireland Multiple Deprivation Measures 2017: Technical Report. URL: https://www.nisra.gov.uk/statistics/deprivation/northern-ireland-multiple-deprivation-measure-2017-nimdm2017.

Pamuk, E.R., 1985. Social class inequality in mortality from 1921 to 1972 in England and Wales. Popul. Stud. (NY). 39, 17–31. doi:10.1080/0032472031000141256.

Public Health Scotland,. Population estimates. URL: https://www.opendata.nhs.scot/dataset/population-estimates/.

R Core Team, 2021. R: A Language and Environment for Statistical Computing. R Foundation for Statistical Computing. Vienna, Austria. URL: https://www.R-project.org/.

Ralston, K., Dundas, R., Leyland, A.H., 2014. A comparison of the Scottish Index of Multiple Deprivation (SIMD) 2004 with the 2009 + 1 SIMD: does choice of measure affect the interpretation of inequality in mortality? Int. J. Health Geogr. 13, 27. doi:10.1186/1476-072X-13-27.

Regidor, E., 2004. Measures of health inequalities: Part 2. J. Epidemiology Community Health 58, 900–903. doi:10.1136/jech.2004.023036.

Revelle, W., 2022. psych: Procedures for Psychological, Psychometric, and Personality Research. Northwestern University. Evanston, Illinois. URL: https://CRAN.R-project.org/package=psych. r package version 2.2.5.

Salmond, C.E., Crampton, P., 2012. Development of New Zealand’s deprivation index (NZDep) and its uptake as a national policy tool. Can. J. Public Heal. 103, S7–11.

Schederecker, F., Kurz, C., Fairburn, J., Maier, W., 2019. Do alternative weighting approaches for an Index of Multiple Deprivation change the association with mortality? A sensitivity analysis from Germany. BMJ Open 9, 1–9.

Schiøtz, M.L., Stockmarr, A., Høst, D., Glümer, C., Frølich, A., 2017. Social disparities in the prevalence of multimorbidity - a register-based population study. BMC Public Health 17. doi:10.1186/S12889-017-4314-8.

Scottish Government, 2012. Scottish Index of Multiple deprivation 2012: A National Statistics Publication for Scotland.

Scottish Government, 2020. SIMD 2020 technical notes. URL: https://www.gov.scot/publications/simd-2020-technical-notes/.

Scottish Government, 2022. Long-term Monitoring of Health Inequalities. URL: https://www.gov.scot/binaries/content/documents/govscot/publications/statistics/2022/03/long-term-monitoring-health-inequalities-march-2022-report2/documents/long-term-monitoring-health-inequalities/long-term-monitoring-health-inequalities/govscot{%}3Adocument/long-term-monitoring-health-inequalities.pdf.

Scottish Public Health Observatory,. Measuring health inequalities. URL: https://www.scotpho.org.uk/comparative-health/measuring-inequalities.

Strömberg, U., Baigi, A., Holmén, A., Parkes, B.L., Bonander, C., Piel, F.B., 2021. A comparison of small-area deprivation indicators for public-health surveillance in Sweden. Scand. J. Public Health, 140349482110303doi:10.1177/14034948211030353.

Thompson, L.C., Wilson, P.M.J., McConnachie, A., 2013. A universal 30-month child health assessment focussed on social and emotional development. J. Nurs. Educ. Pract. 3, 13–22. doi:10.5430/jnep.v3n1p13.

Townsend, P., 1987. Deprivation. J. Soc. Policy 16, 125–146. doi:10.1017/S0047279400020341.

Watson, V., Dibben, C., Cox, M., Atherton, I., Sutton, M., Ryan, M., 2019. Testing the Expert Based Weights Used in the UK’s Index of Multiple Deprivation (IMD) Against Three Preference-Based Methods. Soc. Indic. Res. 144, 1055–1074. doi:10.1007/s11205-018-02054-z.

Welsh Government, 2019. Welsh Index of Multiple Deprivation (WIMD) 2019 Technical report. URL: https://gov.wales/sites/default/files/statistics-and-research/2019-12/welsh-index-multiple-deprivation-2019-technical-report.pdf.

Wickham, H., Averick, M., Bryan, J., Chang, W., McGowan, L.D., François, R., Grolemund, G., Hayes, A., Henry, L., Hester, J., Kuhn, M., Pedersen, T.L., Miller, E., Bache, S.M., Müller, K., Ooms, J., Robinson, D., Seidel, D.P., Spinu, V., Takahashi, K., Vaughan, D., Wilke, C., Woo, K., Yutani, H., 2019. Welcome to the tidyverse. J. Open Source Softw. 4, 1686. doi:10.21105/joss.01686.

